# A machine learning approach to predict mortality and pulmonary hypertension severity in newborns with congenital diaphragmatic hernia

**DOI:** 10.1101/2024.07.25.24311009

**Authors:** Luana Conte, Ilaria Amodeo, Giorgio De Nunzio, Genny Raffaeli, Irene Borzani, Nicola Persico, Alice Griggio, Giuseppe Como, Mariarosa Colnaghi, Monica Fumagalli, Donato Cascio, Giacomo Cavallaro

**Author notes:** Corresponding Author. Giorgio De Nunzio. Co-First Author: Luana Conte and Ilaria Amodeo. Co-Last Author: Donato Cascio and Giacomo Cavallaro.

## Abstract

Prenatal prediction of postnatal outcomes in newborns with congenital diaphragmatic hernia (CDH) remains challenging, especially for mortality and neonatal persistent pulmonary hypertension (PPHN). Despite the increasing utilization of advanced artificial intelligence (AI) technologies in the neonatal field, this study is pioneering in exploring AI methodologies in the context of CDH. It represents an initial attempt to implement a Machine Learning (ML) system to predict postnatal mortality and PPHN severity, using prenatal and early postnatal data as input variables. We enrolled 50 patients with isolated left-sided CDH from singleton pregnancies and retrospectively collected clinical and imaging variables from fetal ultrasound (US) and shape features extracted from magnetic resonance imaging (MRI), combined with gestational age and birth weight. A supervised ML model for predicting mortality and PPHN severity was developed, achieving good accuracy (88% for mortality prediction and 82% for PPHN) and sensitivity (95% for mortality and 85% for PPHN). The area under the curve (AUC) of the ROC curve was 0.88 for mortality and 0.82 for PPHN predictions. Our results may lead to novel AI applications in the neonatal field, focusing on predicting postnatal outcomes based on prenatal data, ultimately improving prognostic assessments and intervention strategies for such a complex disease.

**Clinical Trial Registration:** The trial was registered at ClinicalTrials.gov with Identifier NCT04609163

**What is Known:** prenatal prediction of postnatal mortality and severity of pulmonary hypertension in CDH newborns remains challenging and largely based on imaging through the volumetric assessment of fetal lungs.

**What is New:** developing a ML system for predicting PPHN severity and mortality risk based on the integrated assessment of prenatal and early postnatal variables is feasible, with good accuracy.

## INTRODUCTION

Congenital diaphragmatic hernia (CDH) is a rare congenital anomaly characterized by incomplete closure of the diaphragm and herniation of abdominal organs into the chest, resulting in pulmonary hypoplasia, neonatal persistent pulmonary hypertension, and cardiac dysfunction [1–3]. CDH occurs in nearly 1 in 2500 births. Several factors influence the prognosis, such as defect size and location, associated anomalies, presence of liver up in the thorax, and gestational age at birth [4,5]. Risk stratification is essential to identify patients who might benefit from specific interventions and to enable a risk-adjusted analysis of outcomes, healthcare costs, and management approaches. Prenatal and postnatal CDH predicting tools have largely increased and have been validated during the last years based on clinical and instrumental data [6,7,16–18,8–15]. However, a universal risk stratification method has not been identified yet, and an agreed-upon set of risk-specific management guidelines is still lacking [19].

In particular, predicting the severity of Neonatal Persistent Pulmonary Hypertension (PPHN) using conventional prenatal diagnostic methods remains challenging. As a result, there is growing interest in leveraging advanced technologies that favor a timely and accurate prognosis.

Artificial Intelligence (AI) is increasingly applied in the neonatal field to support medical data analysis. Predictive algorithms are being developed using traditional Machine Learning (ML) approaches as well as its more advanced Deep Learning (DL) extension. ML and DL can process and analyze medical data, including images from different sources such as ultrasound, magnetic resonance imaging (MRI), and X-ray. Integrating these algorithms into healthcare systems holds promise for enhancing diagnostic accuracy and disease pattern classification. These algorithms could help predict specific outcomes, guide interventions, and improve the overall quality of care [20,21,30,22–29].

However, to our knowledge, these methodologies still need to be successfully applied to newborns with CDH. The aim of our study was to provide a predictive algorithm for mortality and PPHN in CDH based on the integrated analysis of prenatal and early postnatal data.

## MATERIALS AND METHODS

### Study design

This study represents an exploratory secondary analysis of a retrospective cohort study performed at Fondazione IRCCS Ca’ Granda Ospedale Maggiore Policlinico, Milan, Italy (CLANNISH, Clinical Trials identification n°: NCT04609163) [29]. The study involved the following services: the Fetal Surgery Center, Pediatric Radiology Service, and Neonatal Intensive Care Unit (NICU). Moreover, the Department of Mathematics and Physics at the Università del Salento (Lecce, Italy) and the Department of Physics and Chemistry at the Università degli Studi di Palermo (Palermo, Italy) developed the AI algorithms.

The current study adhered to the principles of good clinical practice and followed the guidelines of the Helsinki Declaration. It received approval from the local ethics committee (Milan Area 2, Italy) with approval number/ID 800_2020bis. However, considering its retrospective design, the ethics committee waived the need for informed consent. The study was also registered on ClinicalTrials.gov with the identifier NCT04609163.

### Patients

The study population, inclusion-exclusion criteria, and a comprehensive description of the primary study design were previously published and are briefly summarized here [29]. Inborn CDH patients born between 01/01/2016 and 30/04/2020 admitted to the NICU at birth were included. The take- charge of the mothers took place at our Fetal Surgery Center at a gestational age of 30+6 weeks or below. Non-isolated CDH and twin pregnancies were excluded. Only left-sided CDH were considered because of their larger numerosity, homogeneity, and variability in liver position, leaving out right-sided CDH.

A total of 50 patients were included in the final study population.

### Data Collection

Clinical maternal and fetal prenatal variables were retrospectively collected using Astraia software (Astraia Software GmbH, Ismaning, Germany) and NeoCare software (GPI SpA, Trento, Italy). A prenatal ultrasound (US) performed between 25+0 and 30+6 weeks of gestation was considered for each patient. In the case of fetal endoscopic tracheal occlusion (FETO), the fetal US was performed before the fetal procedure. Additionally, native sequences from fetal MRI were gathered for 36 out of 50 cases, with separate acquisitions for the lung and liver. The imaging software employed for this study was Synapse PACS and Synapse 3D (FUJIFILM Medical Systems, Lexington, MA, US). Lung volumes were computed using T2 HASTE sequences, selecting the best-quality image plane without motion-induced artifacts [31]. On the other hand, liver volumes were calculated based on T1 VIBE sequences [32]. An experienced pediatric radiologist (IB) freehand delineated Regions Of Interest (ROIs) to define the areas of the left and right lungs and the liver, excluding the pulmonary hila and mediastinal structures for each slice. Organ volumes were calculated using the software. Subsequently, the DICOM (Digital Imaging and Communications in Medicine) files were anonymized and converted to the NIfTI (neuroimaging informatics technology initiative) format for easy manipulation.

### Clinical and Imaging variables

A detailed summary of the clinical and imaging variables included is reported in Table 1.

**Table 1.** Clinical and imaging variables.

| **Table 1. Clinical and imaging variables** |
| --- |
| ***Maternal data***<br>Mother age<br>Ethnicity<br>Number of gestations ending the outcome for fetuses with an Assisted Reproduction (MAR):<br>yes/no<br>Antenatal use of corticosteroids: yes/no<br>Premature rupture of membranes (pPROM): yes/no<br>Gestational age at pPROM<br>Gestational age at CDH diagnosis |
| ***Fetal ultrasound data***<br>Gestational age<br>Estimated Fetal Weight the (EFW)<br>Amniotic Fluid (AF)<br>Umbilical Artery Pulsatility Index |
| Pulmonary Flow Pulsatility Index<br>Pulmonary flow Peak Systolic Velocity<br>Peak early diastolic reversed flow<br>Organ herniation of Bowel, Stomach, Liver: yes/no<br>Observed/expected lung-to-head ratio (tracing method)<br>CDH severity: mild, moderate, severe<br>Fetal Endoscopic Tracheal Occlusion (FETO): yes/no<br>Gestational age at balloon insertion and removal |
| ***MRI data***<br>Gestational age<br>Apparent diffusion coefficient (ADC)<br>Right and left lung volume<br>Total fetal lung volume (TFLV)<br>Observed/expected TFLV (O/E TFLV%)<br>Total liver volume<br>Herniated Liver volume<br>Percentage of liver herniation (%LH)<br>Mediastinal shift angle (MSA)<br>Apparent diffusion coefficient (ADC) of the left and right lung |
| ***Early postnatal data***<br>Gestational age at birth<br>Birth weight |

### Radiomics features

In addition to the clinical variables, standard 3D radiomics features were extracted from the segmented ROIs in the MRI using the freely available and open-source Pyradiomics v. 3.01 software tool [30,33]. Pyradiomics produces many variables, with and without preprocessing by various filters and optional reslicing, with different interpolators. Only features from the original images without preprocessing were considered in this work. Due to significant dissimilarity in the gray-level content of the MRI scans, only shape features were utilized (MeshVolume, VoxelVolume, SurfaceArea, SurfaceVolumeRatio, Sphericity, Maximum3DDiameter, MajorAxisLength, MinorAxisLength, LeastAxisLength, Elongation, and Flatness). The geometric meaning of each feature is detailed in Table 2.

**Table 2.** Geometric meaning of Pyradiomics shape features.

| **Table 2. Geometric meaning of Pyradiomics shape features** |  |
| --- | --- |
| **Feature** | **Geometric Meaning** |
| MeshVolume | The shape volume, calculated using a mesh representation. It is the three-dimensional space enclosed by the surface of the object. |
| VoxelVolume | The volume calculated by counting the number of voxels (3D pixels) within the shape and multiplying by the volume of a single voxel. This represents the discretized volume of the object. |
| SurfaceArea | The total area of the surface of the shape. This quantifies the two-dimensional extent of the object's surface. |
| SurfaceVolumeRatio | The ratio of surface area to volume. This measure indicates how 'compact' an object is; lower values suggest a more compact shape. |
| Sphericity | A measure of how spherical (round) the object is. Perfect spheres have a sphericity of 1. Lower values indicate less spherical shapes. |
| Maximum3DDiameter | The largest distance between any two points on the surface of the shape. This is the maximum length of the object in any dimension. |
| MajorAxisLength | The length of the major axis of the shape, which is the longest dimension in the principal component analysis (PCA) of the object. |
| MinorAxisLength | The length of the minor axis, which is perpendicular to the major axis and is the second longest dimension in the PCA of the object. |
| LeastAxisLength | The shortest axis length from the PCA of the object. It's perpendicular to both the major and minor axes. |
| Elongation | The ratio of the minor axis length to the major axis length. It indicates how much longer the shape is in one direction compared to the other. |
| Flatness | The ratio of the least axis length to the major axis length. This measure indicates how 'flat' or 'elongated' an object is compared to being spherical. |

Variables computed from the gray levels were discarded, avoiding additional image manipulation, such as intensity standardization. A total of 80 features were considered: 56 prenatal variables, 2 very early postnatal variables as gestational age and birth weight, and 22 MRI-extracted shape features (11 from the lungs, 11 from the liver).

To ensure fairness in the classification process, the features were normalized to the 0-1 range using min-max normalization on the training set. The same normalization parameters were then applied to the validation set samples. However, in some cases, the lack of MRI data resulted in missing values in the features extracted by Pyradiomics. This was also observed for non-radiomics features based on the patient’s diagnostic pathway. Imputation by a weighted average was considered [34] to handle these missing values, as in Equation 1:

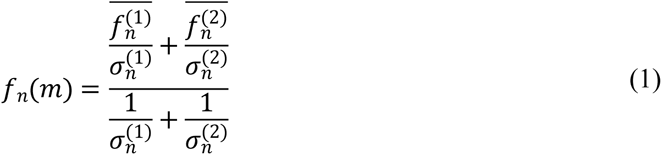

where *f*_*n*_(*m*) is the value to be assigned to the (missing) *n*-th feature for the *m*-th sample, 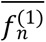 and 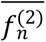 are the average values of the *n*-th features for classes 1 and 2, respectively, and 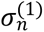 and 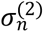 are the corresponding standard deviations. This way, in the approximation of Gaussian distributions, a neutral value for the distributions of the two classes is used as the missing feature.

### Target variables: neonatal persistent pulmonary hypertension and mortality

For each included patient, a neonatologist performed a systematic revision of the first available echocardiogram within 24 hours after birth, focusing on direct and indirect signs of pulmonary hypertension. Data collection was focused on the presence and characteristics of the shunts through patent ductus arteriosus and *foramen ovale*, the characteristics of the intraventricular sept, the estimation of the systolic pulmonary artery pressure through tricuspid valve regurgitation, the systemic pressures, and concomitant use of pulmonary vasodilators. Patients were then stratified according to the presence and severity of pulmonary hypertension into two categories: severe (over- systemic, considered as the positive class) *vs* moderate/mild (iso/under-systemic). According to mortality, the study population was divided into non-survivors (positive class) *vs* survivors.

### Feature selection

We employed the Recursive Feature Elimination (RFE) technique with Cross-Validation (CV), specifically using the Leave One Patient Out CV (LOPO-CV) scheme. The RFE method starts with the entire feature set and recursively removes the minor essential features based on a chosen metric (in this case, accuracy) until the desired number of features is reached. Typically, the final number of features to select is a parameter that needs to be specified. This parameter was determined dynamically by varying its value and calculating the corresponding accuracy in our approach. We then selected the parameter value that maximized accuracy. The Random Forest (RF) classifier was used to evaluate the various configurations.

### Training

To exploit the available samples as much as possible, we used a LOPO-CV scheme. In this method, we selected one patient as the validation set while using the remaining patients for training. We trained several classifiers and obtained performance metrics such as the confusion matrix, sensitivity, specificity, area under the ROC curve (AUC), and the area under the P-R curve. All the optimization steps were based on maximizing accuracy.

### Classifiers

Three classification algorithms were tested: eXtreme Gradient Boosting (XGBoost), Support Vector Machine (SVM), and K-Nearest Neighbors (KNN). The first classifier was used because it natively and effectively deals with missing clinical values. The choice of the other two classifiers was due to their ability to allow good performance in conditions of a limited number of available samples, as they are characterized by a reduced number of parameters to be tuned [35]. Hyperparameter tuning was performed to avoid overfitting and improve model performance.

## RESULTS

The final study population consisted of 50 patients: 26 severe (52%) and 24 moderate/mild (48%) cases of PPHN. According to mortality, 37 survivors (74%) and 13 non-survivors (26%) were present.

As regards mortality analysis, the feature selection procedure led to the choice of 10 out of 80 features, in particular: maternal age, gestational age at CDH diagnosis, CDH severity (mild, moderate, severe), centile of estimated fetal weight (EFW), observed/expected lung to head ratio (o/e LHR) by tracing method, umbilical artery pulsatility index, left and right fetal lung volume (FLV) at MRI, observed/expected total fetal lung volume (o/e TFLV), gestational age at birth, and birth weight.

Remarkably, no shape features were selected. Table 3 shows the classification results obtained for mortality prediction.

**Table 3.**
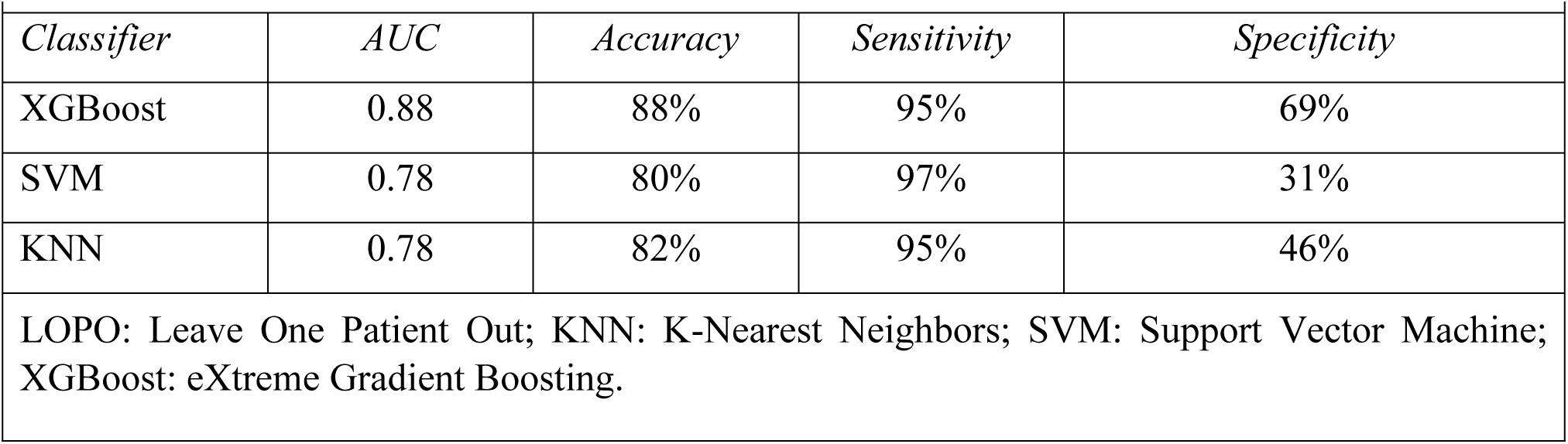
Classification figures of merit for mortality, computed in a LOPO scheme.

|  |  |  |  |  |
| --- | --- | --- | --- | --- |
| *Classifier* | *AUC* | *Accuracy* | *Sensitivity* | *Specificity* |
| XGBoost | 0.88 | 88% | 95% | 69% |
| SVM | 0.78 | 80% | 97% | 31% |
| KNN | 0.78 | 82% | 95% | 46% |
| LOPO: Leave One Patient Out; KNN: K-Nearest Neighbors; SVM: Support Vector Machine; XGBoost: eXtreme Gradient Boosting. |  |  |  |  |

The results of the different classification methods were comparable regarding AUC and accuracy, though XGboost had better performance. Figure 1 shows the ROC and P-R curves for mortality prediction obtained by XGboost with feature selection. The trained model correctly identified 88% of cases and achieved a sensitivity of 95% and a specificity of 69%. The AUC from the ROC curve was 0.87, while the P-R curve subtended an area of 0.95 (with the frequency of positive cases equal to 52%). From the P-R curve, precision drops after 50% sensitivity but remains more than 85% when sensitivity is 90%, and even if we require complete sensitivity, precision remains relatively high (around 80%).

**Fig 1.**
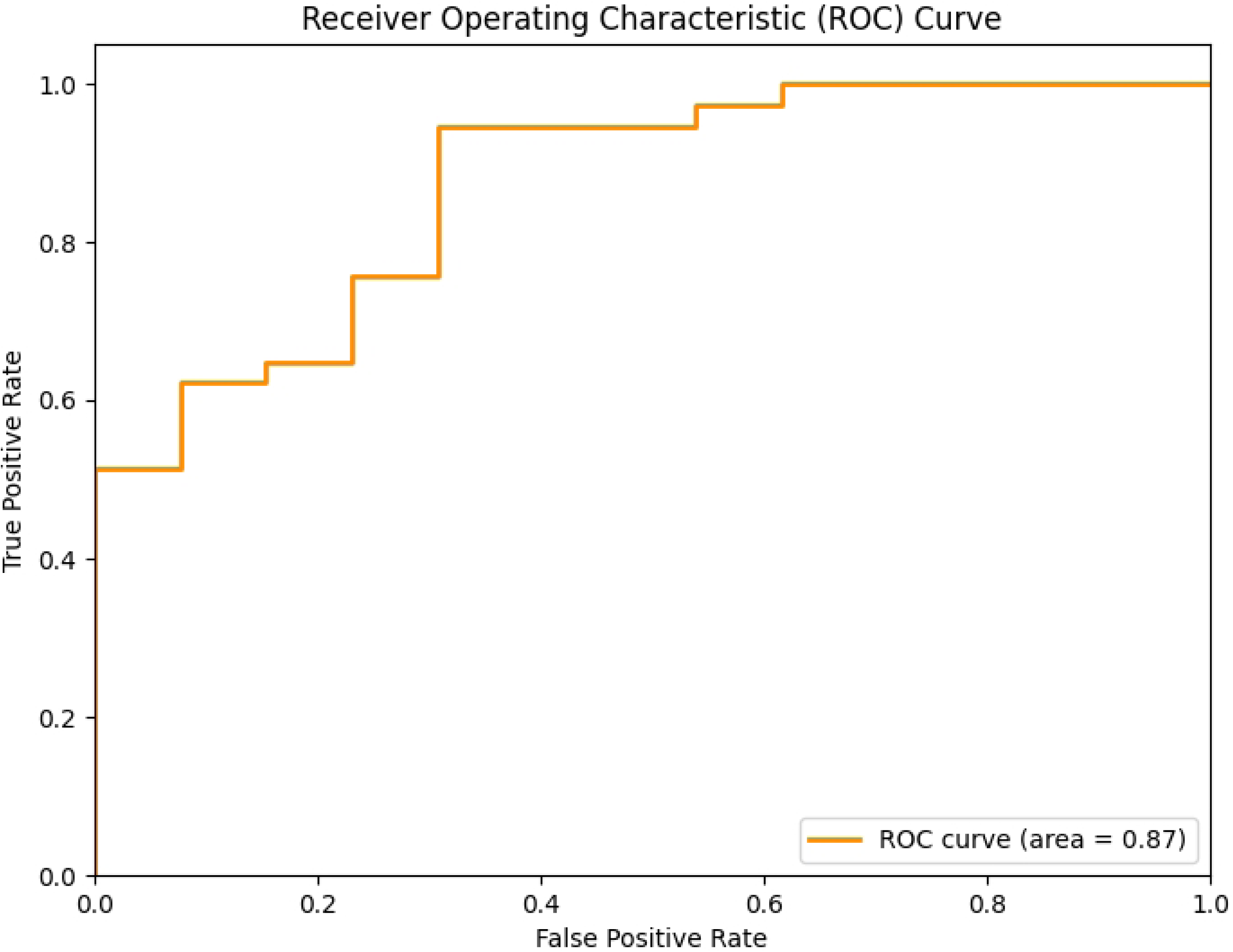
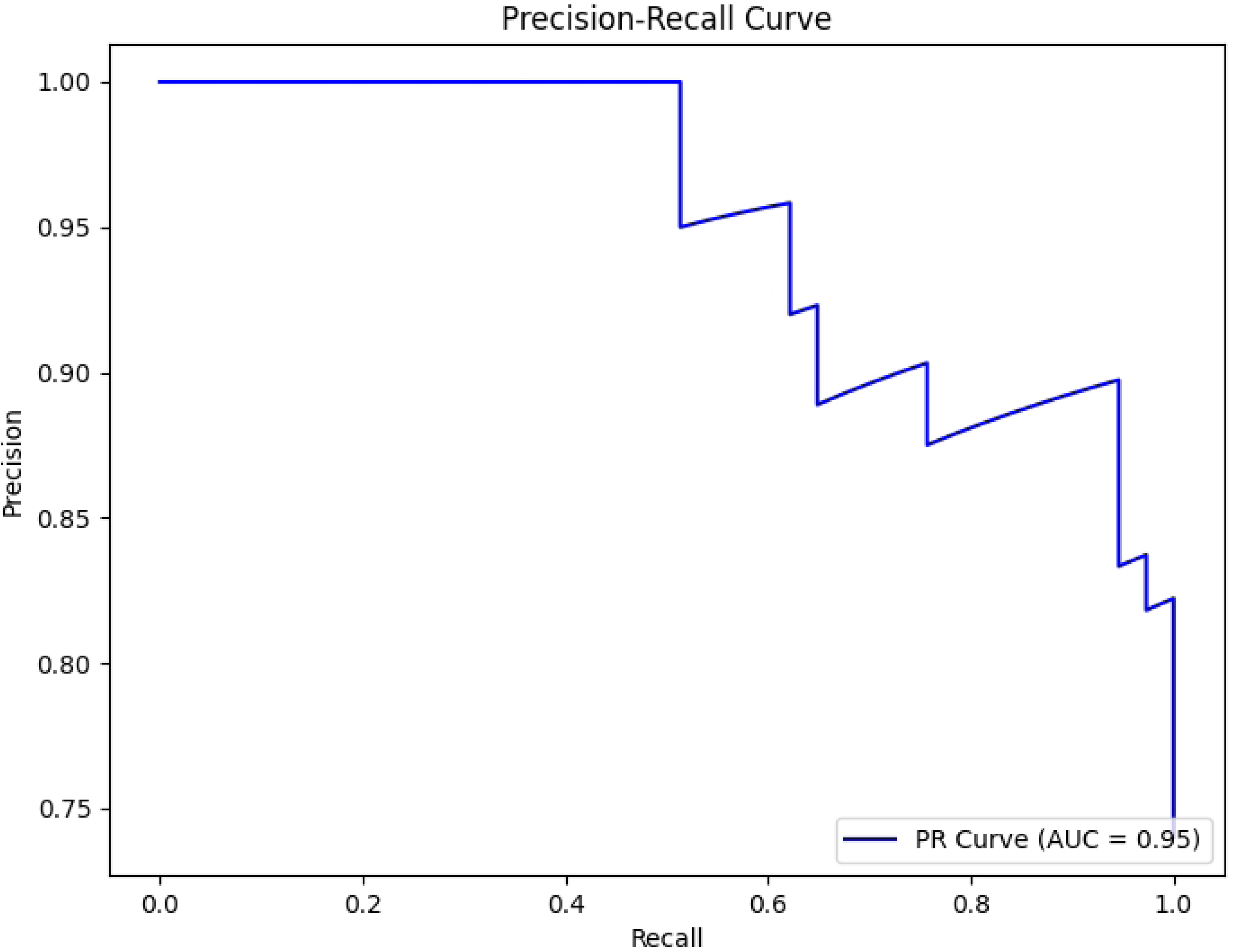
Mortality prediction. Left: ROC curve, right: P-R curve, obtained with the XGBoost classifier on prenatal clinical variables. No shape features extracted from MRIs were used, as required by the feature selection procedure.

As far as PH is concerned, feature selection led to the identification of the 14 features, in particular: gestational age at CDH diagnosis, liver position, grading of stomach herniation, gestational age at US ultrasound, centile of EFW, umbilical artery pulsatility index, peak early diastolic reversed flow, o/e TFLV, apparent diffusion coefficient of left lung, original shape elongation, gestational age at birth, and birth weight.

In this case, both clinical and shape features were selected. Table 4 reports the classification results obtained for PPHN classification. The results produced by the different classification methods show that XGboost also performed better in this case.

**Table 4.** Classification figures of merit for PPHN, computed in a LOPO scheme.

|  |  |  |  |  |
| --- | --- | --- | --- | --- |
| **Classifier** | **AUC** | **Accuracy** | **Sensitivity** | **Specificity** |
| XGBoost | 0.82 | 82% | 85% | 79 |
| SVM | 0.75 | 74 | 79 | 71 |
| KNN | 0.65 | 54 | 53 | 33 |
| LOPO: Leave One Patient Out; KNN: K-Nearest Neighbors; PPHN: Neonatal Persistent Pulmonary Hypertension; SVM: Support Vector Machine; XGBoost: eXtreme Gradient Boosting. |  |  |  |  |

The XGboost classifier demonstrated significantly superior classification capabilities to the other two classifiers (Tables 3 and 4).

Figure 2 shows the ROC and P-R curves for PPHN classification obtained by XGboost with the features selection. The AUC from the test ROC curve was 0.82, while the P-R curve subtended an area of 0.75 (with the frequency of positive cases equal to 26%). The trained model correctly identified 82% of cases and achieved a sensitivity of 85% and a specificity of 79%. From the P-R curve, we deduce that even at very high sensitivity values (about 83-84%), precision is more than 80%.

**Fig 2.**
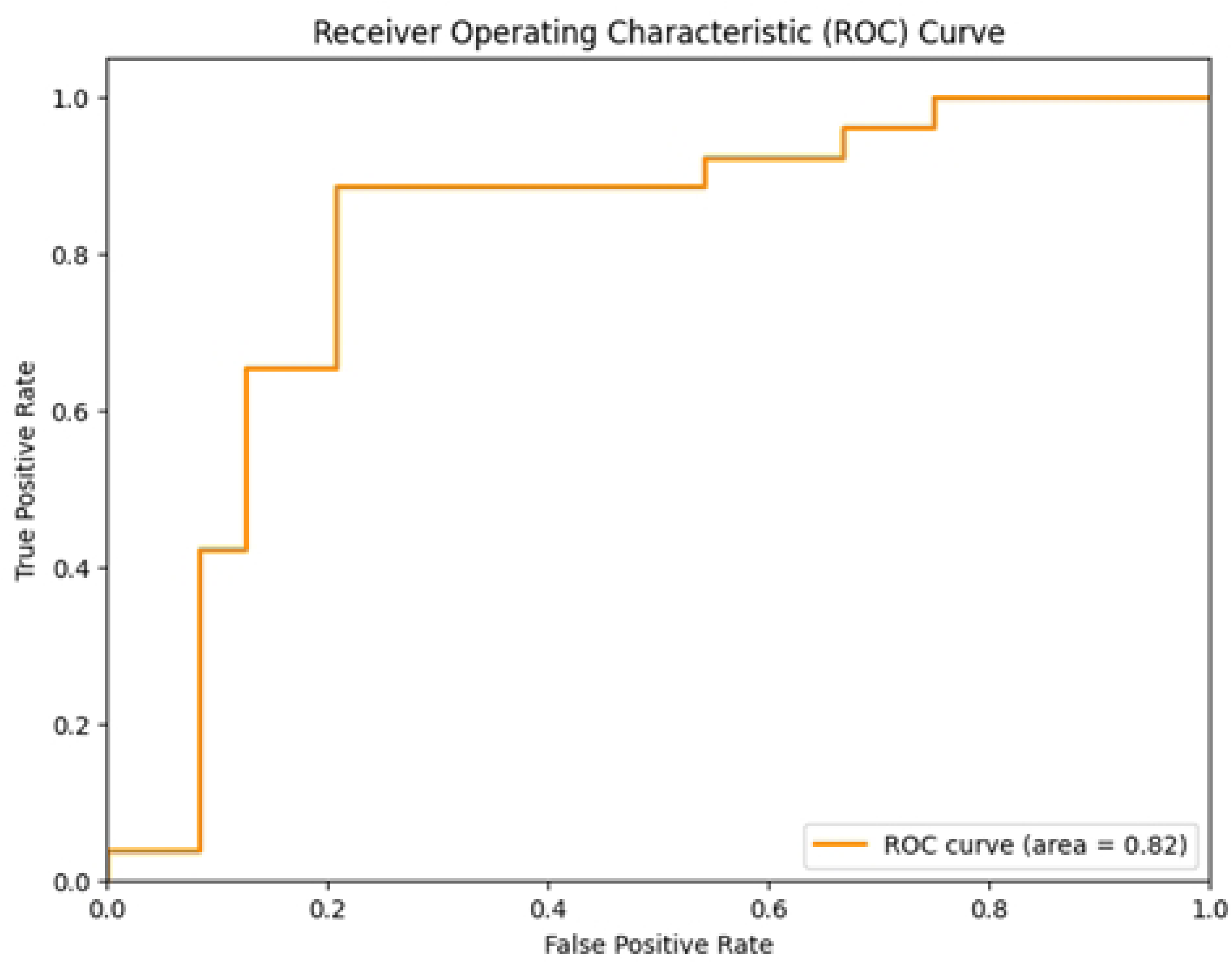
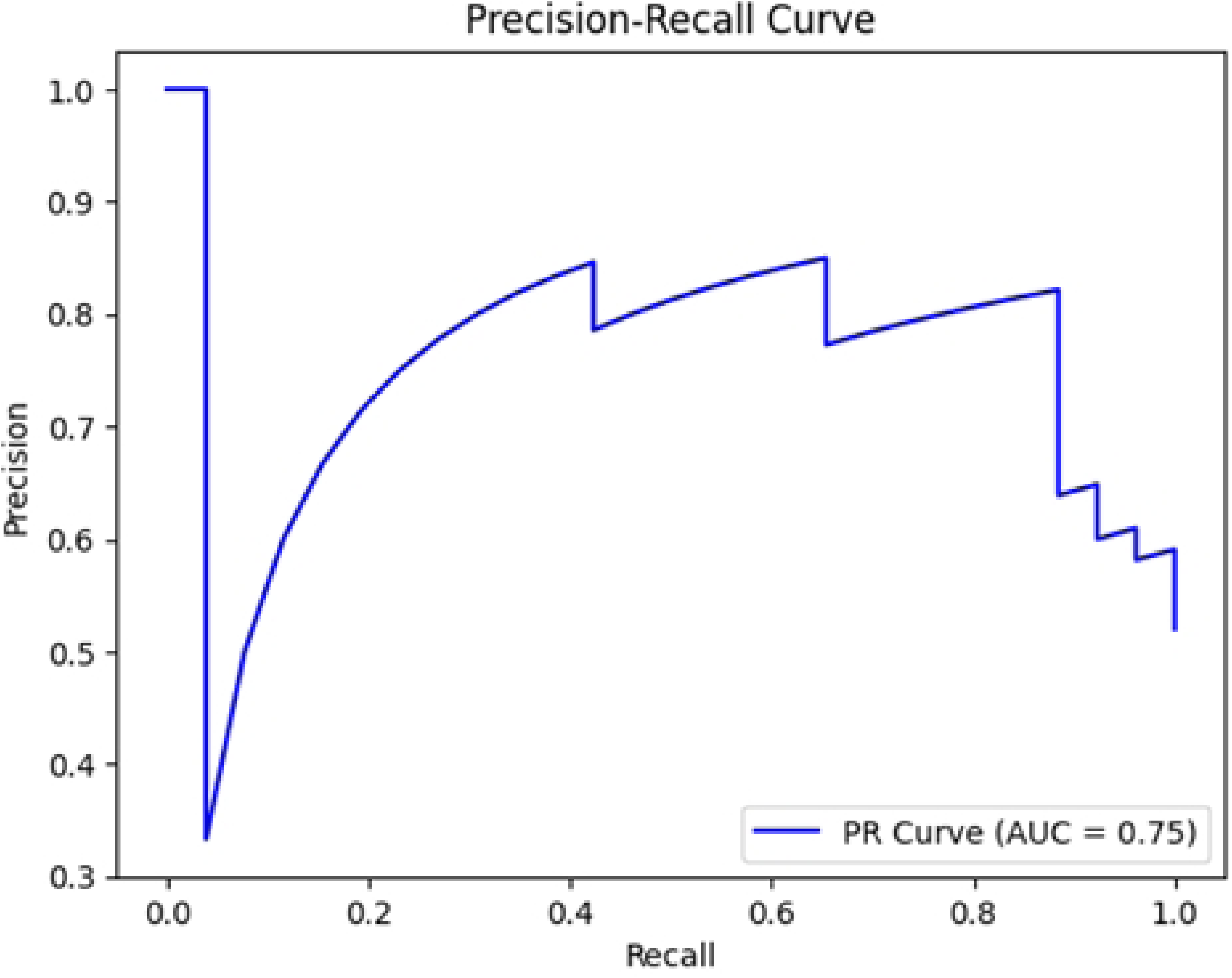
Left: the ROC curve; right: the Precision-Recall (P-R) curv**e** for PPHN predictions with the XGBoost classifier on prenatal clinical variables and shape features extracted from MRIs.

## DISCUSSION

Congenital diaphragmatic hernia (CDH) is a life-threatening anomaly requiring high-skilled and multidisciplinary team of experts for appropriate management since from antenatal diagnosis [36]. Despite advancements over time, morbidity and mortality remain significant (20–40%), even within high-volume tertiary referral centers [37–39]. An estimated quarter of survivors experience neurodevelopmental impairments across all domains, encompassing motor and sensory (hearing, visual) deficits as well as cognitive, language, and behavioral impairments [40].

CDH patients exhibit varying degrees of pulmonary hypoplasia and abnormal pulmonary vascular disease, resulting in varying extents of pulmonary hypertension. Up to 30–40% of newborns with CDH experience concomitant cardiac ventricular dysfunction [41,42]. PPHN is associated with adverse outcomes in CDH patients, underscoring the critical nature of its management in the care of these infants [43].

Various clinical and laboratory parameters and prognostic indices in the perinatal period have been subject to study to predict postnatal outcomes [39,44–47]. The identification of variables predictive of mortality is paramount for clinical decision-making and parental guidance. The o/e LHR and o/e TFLV serve as pivotal metrics in this regard. Each of these parameters evaluates the extent of pulmonary hypoplasia associated with CDH, a critical determinant of both survival and long-term prognosis.

The o/e LHR has been widely studied and utilized in the prediction of postnatal survival in cases of isolated CDH. Jani et al. highlighted the significance of the o/e LHR in predicting survival in fetuses with isolated diaphragmatic hernia [48]. Snoek et al. further assessed the predictive value of the o/e LHR for survival and chronic lung disease (CLD) in survivors with left-sided CDH, reflecting its ongoing relevance in an era of standardized neonatal treatment [49]. Their multicenter study underscores the evolving understanding of o/e LHR in predicting outcomes for CDH patients.

On the other hand, the o/e TFLV exhibits a stronger correlation with postnatal outcomes than the absolute lung volume. Moreover, a growing body of evidence supports the superior accuracy of o/e TFLV in predicting survival compared to ultrasound-based estimations of lung size, which may not fully account for the ipsilateral lung and could thus underestimate the effective lung volume [50–55]. In cases of isolated CDH, o/e TFLV has demonstrated efficacy in distinguishing survival, with an o/e TFLV < 25% being associated with more severe forms and a reduced survival rate [54,56–60]. Furthermore, o/e TFLV has been shown to forecast the necessity for extracorporeal membrane oxygenation (ECMO) after birth, with the combined assessment of lung volumetry and o/e LHR proving more effective than ultrasound alone in predicting the need for ECMO [61–64].

Prenatal prediction of PPHN plays a crucial role in prenatal management, delivery planning and postnatal care. However, while both o/e LHR and o/e TFLV offer insights into the extent of CDH-associated pulmonary hypoplasia, their predictive value for PPHN necessitates careful consideration. Our findings support the possibility of successfully developing a ML system for predicting PPHN severity and mortality risk based on the integrated assessment of prenatal and early postnatal variables.

To achieve our goal, we enrolled 50 left-sided CDH cases. The dataset was relatively balanced concerning PPHN, with 26 severe and 24 moderate/mild cases, whereas mortality classes included 37 survivors and 13 non-survivors. We combined prenatal clinical and imaging data with gestational age and weight at birth, which both play a key role in survival in neonatal patients, especially those in critical conditions. In addition, standard 3D radiomics features were extracted from the segmented ROIs using the freely available Pyradiomics software tool. This software package facilitated automatic reslicing with a selected interpolator and computed multiple radiomics variables. As the MRIs exhibited significant variations in grayscales, which would have required some form of intensity standardization to use features based on gray values, we only utilized shape features to avoid additional image manipulation and discarded variables based on the gray levels.

A feature selection phase was executed for both postnatal target variables, mortality, and PPHN. The RFE approach used RF classification to evaluate different configurations, which was appropriate for several reasons. First, this approach provides features of relative importance during the training process. Each time a decision tree is constructed, the model tracks how much each feature contributes to reducing the cost function, usually Gini impurity or entropy. An importance score for each feature is obtained by averaging this importance across all trees. Second, because of its "forest" nature, a RF is robust and less prone to overfitting than individual decision trees. This means that the computed importance of features is more reliable and less affected by noise in the data. Finally, RFs can handle highly correlated features without special preprocessing. In the presence of correlations, this approach can distribute importance among correlated features, providing a complete picture of each feature contribution.

We conducted a comprehensive evaluation of three classification algorithms: XGBoost, SVM, and KNN. Our models were trained using prenatal and early postnatal clinical variables, as well as selected shape features extracted from MRI data. Interestingly, we discovered that XGBoost outperformed the other models and emerged as the best classification model for both clinical targets. The supervised ML models, designed to predict PPHN severity and neonatal mortality, showed promising preliminary results. Our study suggests that predicting mortality and PPHN severity in the prenatal and very early postnatal period can be feasible by ML applications, achieving accuracies of 88% for mortality and 82% for postnatal PPHN. With significant accuracy rates and reliable sensitivity, this model has the potential to revolutionize prognostic assessment in CDH, eventually improving patient outcomes. By implementing the algorithm, risk categories could be simulated based on available prenatal data and assuming gestational age and estimated fetal weight at birth. The algorithm could also be updated in real time at subsequent obstetric visits or based on the threat of preterm delivery, as prematurity plays a significant role in survival, especially in infants with underlying disease. This would assist with parenting counseling, birth planning, and postnatal care. To the best of our knowledge, our studies are the first to explore the application of AI methods to CDH [29,30].

Despite being encouraging, some limitations must be considered. First, the restricted dataset deriving from the rarity of the condition represented the weakest point. An appropriate number of cases during training/validation and data interpretation is crucial for ML applications. Potential strategies may involve collaborating with other institutions and prospectively considering including future cases to augment the study population. Another critical aspect is data inhomogeneity, specifically the lack of a standard grayscale in the images. This would require a standardization procedure, after which gray- level-based features could be used to increase ML quality for classification purposes. Nonetheless, MRI standardization is a delicate process that involves profound changes in image gray levels, which might even make ML procedures less accurate. Consequently, we preferred to simply discard ML features based on the gray-level content of ROIs, only using shape features. Interestingly, no shape features were selected for the mortality target, whereas clinical and shape variables were chosen for the PPHN target. We can speculate that the information provided by the images is more closely related to the structure and architecture of the lung parenchyma, which directly impacts the disease’s pathophysiology. On the other hand, mortality may be an indirect result of these structural alterations, influenced by many factors. Although a conclusive interpretation is not yet possible, this aspect deserves further investigation, and an increase in the study population and image optimization are crucial. Finally, the retrospective data collection is largely affected by missing or inaccurate data and may be time-consuming for the clinician. Standardized assessment and computerized data collection could improve the dataset quality.

## CONCLUSIONS

Although with limitations, with reasonable accuracy, a ML approach for predicting mortality and PPHN severity of CDH newborns using prenatal and very early post-natal variables appears feasible. Our results could pave the way for new AI applications in the neonatal field. They would enable risk- adjusted analyses of outcomes, healthcare costs, and management strategies, ultimately improving the overall quality of care.

## Data Availability

All relevant data are within the manuscript and its Supporting Information files.

## Statements and Declarations

### Competing Interests

The authors have no relevant financial or non-financial interests to disclose.

### Funding

This study was (partially) funded by the Italian Ministry of Health - Current Research IRCCS.

### Author Contributions

L.C., I.A., G.D.N., G.R., I.B., D.C., and G.C. contributed to the study’s conception and design; L.C., I.A., G.D.N., G.R., I.B., D.C., G.C. (Giuseppe Como), N.P., and G.C. (Giacomo Cavallaro) contributed to the study’s methodology, investigation, and data curation; I.B. contributed to manual segmentation; L.C., G.D.N., L.C., and D.C. contributed to ML and DL analysis; L.C., G.D.N., and D.C. performed the statistical analysis; L.C., I.A., G.D.N., G.R., I.B., D.C., and G.C. (Giacomo Cavallaro) wrote the initial draft preparation of the manuscript; L.C., I.A., G.D.N., G.R., I.B., D.C., G.C. (Giuseppe Como), N.P., M.C., M.F., and G.C. (Giacomo Cavallaro) wrote, reviewed, and edited the manuscript; L.C., G.D.N., and D.C. contributed to designing the figures; G.D.N. and G.C. (Giacomo Cavallaro) contributed equally to the visualization of the manuscript; G.D.N. and G.C. (Giacomo Cavallaro) contributed to the supervision and project administration of the study. All authors have read and agreed to the published version of the manuscript.

### Ethics approval

The present study was conducted using the principles of good clinical practice and the Helsinki Declaration. It was approved by the local ethics committee (Milan Area 2, Italy) with approval number 800_2020bis. However, due to the study’s retrospective nature, the Ethics Committee waived informed consent. The study was registered at ClinicalTrials.gov with the identifier NCT04609163.

### Consent to participate

Written informed consent was obtained from the parents.

### Conflicts of Interest

The authors declare that the research was conducted without any commercial or financial relationships that could be construed as a potential conflict of interest.

## Notes

### Competing Interest Statement

The authors have declared no competing interest.

### Clinical Trial

The study was registered on ClinicalTrials.gov with the identifier NCT04609163.

### Funding Statement

The author(s) received no specific funding for this work.

### Author Declarations

The current study adhered to the principles of good clinical practice and followed the guidelines of the Helsinki Declaration. It received approval from the local ethics committee (Milan Area 2, Italy) with approval number/ID 800_2020bis. However, considering its retrospective design, the ethics committee waived the need for informed consent.

